# The reproductive index from SEIR model of Covid-19 epidemic in Asean

**DOI:** 10.1101/2020.04.24.20078287

**Authors:** Pongkeaw Udomsamuthirun, Grittichon Chanilkul, Pongkarn Tongkhonburi, Chatcharawan Meesubthong

## Abstract

As we calculate analytic to link the coefficient of third-order polynomial equations from raw data of an Asean to the SEIR model. The Reproductive index depending on the average incubation period and the average infection period and the coefficient polynomial equations fitted from raw are derived. We also consider the difference of the average incubation period as 5 days and 3 days with the average infection period as 10 day of an Asean. We find that the value of *R*_0_ are Indonesia (7.97), Singapore (6.22), Malaysia (3.86), Thailand (2.48), respectively. And we also find that Singapore has 2 values of *R*_0_ as 1.54 (16 Feb to 37 March) and 6.22 (31 March-4 April).The peak of infection rate are not found for Singapore and Indonesia at the time of consideration. The model of external stimulus is added into raw data of Singapore and Indonesia to find the maximum rate of infection. We find that Singapore need more magnitude of external stimulus than Indonesia. And the external stimulus for 14 days can stimulate to occur the peak of infected daily case of both country.

## 1. Introduction

An Asean (The Association of Southeast Asian Nations) is consist of 10 ten member states as Indonesia, Malaysia, Philippines, Singapore, Thailand, Brunei Darussalam, Viet Nam, Lao PDR, Myanmar, and Cambodia. And we try to predict the number of coronavirus (Covid-19) victims as number of persons who caught the infection and got sick only in this area. The complicated mathematical models are necessary for long-time predictions. The SIR (Susceptible-Infectious-Removed) model are used to obtain the prediction values of the model parameters using the statistical approach for predication the number of infected, susceptible and removed persons [1,2]. The Susceptible-Infectious-Recovered/Death (SIRD) Model was used to formulate an optimal control problem with an expanded epidemic model to compute (Non-pharmaceutical) implementation strategy.[3] A modify SIR called SEIRUS model (Susceptible – Exposed – Infectious – Removed – Undetectable – Susceptible) is generated for evaluate the new deterministic pandemic Covid-19 endemic that originally developed for the control of the prevalence of HIV/AIDS in Africa.[4] The Susceptible-Exposed-Infectious-Removed (SEIR) model was adopt to be SEIRNDC [5] that the total population size N with two extra classes “D” mimicking the public perception of risk regarding the number of severe and critical cases and deaths; and “C” representing the number of cumulative cases. This model proposed the compartmental model that sustained human-to-human transmission of Covid-19 after December 2019 of Wuhan, China.

In this research, we use the daily cases and total cases of Covid-19 infection population of an Asean from website: Worldometer [6]: https://www.worldometers.info/coronavirus/ between 15/02/2020 to 15/04/2020. The raw data are fitted with the regression method of third-order polynomial formula. We calculate analytic to link the coefficient of polynomial to the Reproductive index (R_0_) of the SEIR model (Susceptible-Exposed-Infected-Removed).

## 2. Model

We apply the well-known SEIR compartmental model [7] (Susceptible-Exposed-Infected-Removed) for the prediction properties of how a disease spread. The variable description is *S*(*t*) is number of susceptible populations, E(t) is the number of exposed populations, *I*(*t*) is number of infected populations, *R*(*t*) is number of infected populations quarantined and expecting recovery at time. There is no emigration from the total population and there is no immigration into the population. A negligible proportion of individuals move in and out of at a given time that

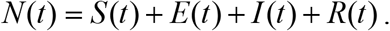

The people susceptible are able to get infected when they contact infectious people. Once infected, they move into the infectious compartment. People recovered are assumed to be immune within our study horizon. Then

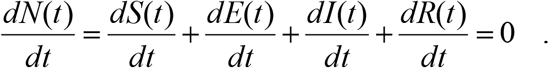

The dynamics non-linear differential equations are below

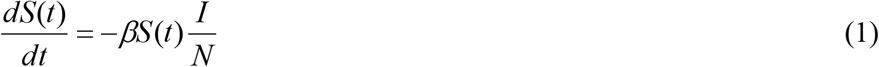

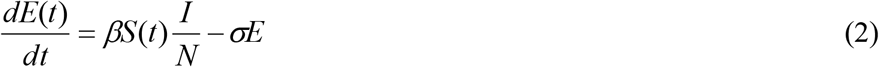

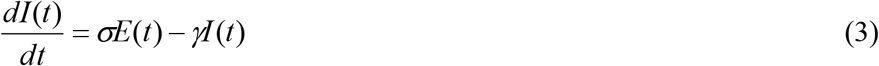

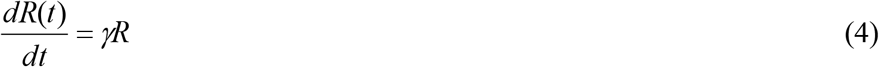

Here, the *β, σ* and *γ* denoted the infection rate, the onset rate, and the removal rate. The 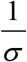 and 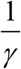 imply the average incubation period and the average infection period. Base on Ref.[8,9], the *σ* = 0.2 and *γ* = 0.1. The normalize parameters of model are defined by

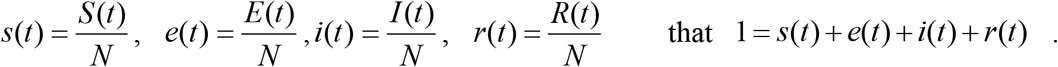

In our model, we assume that the rate of infection is in the form of third-order polynomial formula as

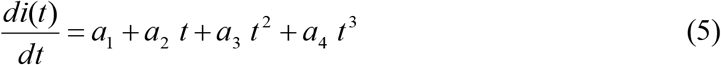

Integrate with respect to time, we can get the total accumulation of infective population that

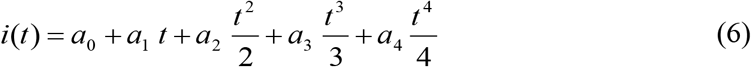

Here *a*_0_ is the total cases found in the day before. *a*_1_, *a*_2_, *a*_3_ *and a*_4_ are constant that found from regression method on the raw data from website: Worldometer at https://www.worldometers.info/coronavirus/.

After take some calculate on solving the first order differential equation on Eqs.(1-6), we get

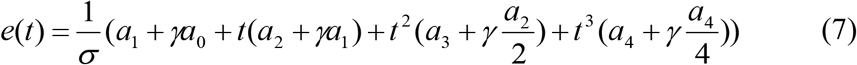

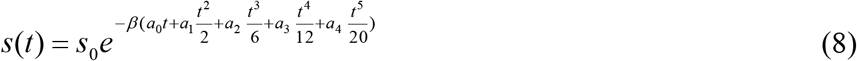

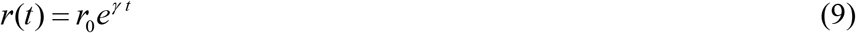

Substitution Eq.(6), Eq.(7) and Eq.(8) in to Eq.(2), we get

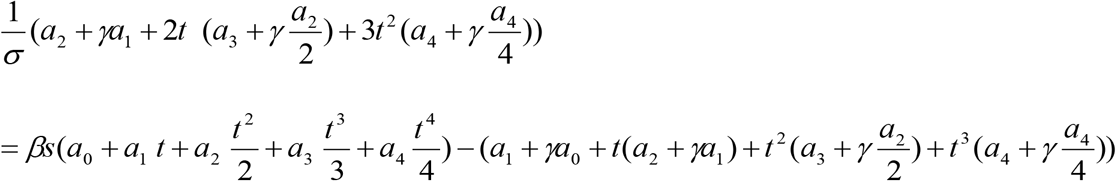

By setting t=0, and 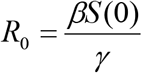, the reproductive index in the parameters of raw data is derived as

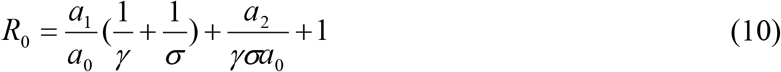

The reproductive index is defined as the dominant eigenvalue of the matric *FV* −1; the linearized infection subsystem F and V. In our model, we assume that there are two disease state but one way to create new infection, *X* = (*e,i*). The linearized infection subsystem F and V are in form 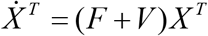 that R_0_ is the dominant eigenvalue of the matrix *FV* ^−1^. From SEIR model, the linearized infection subsystem are 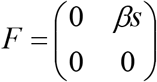 and 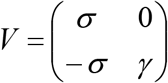. Then, we can get 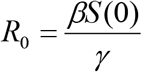. Our R_0_ is agreed with Ref. [10,11]. The reproductive rate or the reproductive index, R_0_ is the course of an epidemic shape. It represents the number of further cases each new case will give rise to. For high value of R_0_, the number of newly infected people climbs more quickly to a maximum than low value of *R*_0_. The higher R_0_, the higher infectious population are.

The World Health Organization (WHO) reported an incubation period for COVID-19 between 1 and 14 days but most commonly around five days. The most infection period is thought to be 1 to 3 days before symptoms start, and in the first 7 days after symptoms begin. Mean incubation period observed are 3.0 days (0 - 24 days range case-study) and 5.2 days (4.1 - 7.0 days range case-study) [6].

Base on Ref.[8] and [9], the *σ* = 0.2 and *γ* = 0.1, or 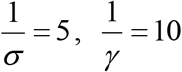. The Reproductive index becomes

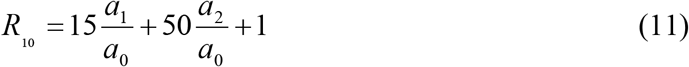

And base on WHO, we set 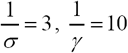, that

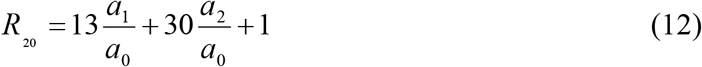

Here *a*_0_ is the accumulation infected population before the date of calculation. *a*_1_ *and a*_2_ are constant values and the coefficient of term “t” of 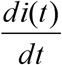, respectively.

## 3. Results and Discussion

When the spreading of viruses in a population occurred, the number of new cases rises rapidly, peaks, and then declines called the epidemiological curve. The spreading curve should be the flatten as the spreading the infections out of time. In this paper, we model the shape of spreading rate to be the third-order polynomial that we called “bell-shape “. According to the total accumulate infected cases, the shape of curve should be gotten the saturation value so that the rate of infectious should be the bell-shape at the end of disease spreading. After analysis the raw data, we find that the daily new cases of Asean are divided into 2 cases; the bell-shape and no bell-shape. The bell-shape cases mean that the infection is in the state of the beginning of the saturated status as we can find the maximum value and we cannot find the maximum value for no bell-shape case. However, both cases can be solve to find the *R*_0_.

An Asean is consist of 10 ten member states as Indonesia, Malaysia, Philippines, Singapore, Thailand, Brunei Darussalam, Viet Nam, Lao PDR and Myanmar, and Cambodia. Because of not enough data to calculate efficiency, we ignore Lao PDR and, Myanmar, Cambodia for our consideration. Here, the raw data fitted well with *R*_*fit*_ ^2^ > 0.80 are used to find the reproductive index so there are only 4 country suitable as Malaysia, Singapore, Thailand and Indonesia.

From Eq.(10), the Reproductive index (*R*_0_) depend on the average incubation period 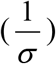 and the average infection period 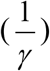. Upon to the difference on the average incubation period as 5 days and 3 days with the average infection period as 10 day, Eq.(10 become to Eq.(11) and Eq.(12) as 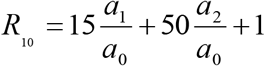 and 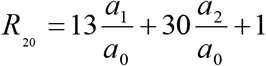. And *a*_0_ is the accumulation infected population before the date of calculation [6]. The others constant, *a*_1_ *and a*_2_, come from our fitting equation. The results are shown in Table 1.

**Table 1.** The Reproductive index (*R*_0_) of Malaysia, Thailand Singapore and Indonesia.

| State | $a_0$ | $a_1$ | $a_2$ | $R_{fit}^2$ | $R_0$ | | |
| --- | --- | --- | --- | --- | --- | --- | --- |
| | | | | | $R_{10}$ | $R_{20}$ | $R_0$ Average |
| Malaysia | 22 | 7.7413 | -5.3826 | 0.81 | 5.96 | 1.77 | 3.86 |
| Thailand | 43 | 9.4464 | -7.0428 | 0.80 | 3.89 | 1.06 | 2.48 |
| Singapore-1 | 72 | 4.385 | -0.5598 | 0.88 | 1.52 | 1.56 | 1.54 |
| Singapore-2 | 147 | 48.206 | 2.3086 | 0.85 | 6.70 | 5.73 | 6.22 |
| Indonesia | 117 | 2.1858 | 19.633 | 0.85 | 9.67 | 6.28 | 7.97 |

As there are only 4 countries that have enough data to fit well with third-order polynomial; Malaysia, Thailand, Singapore, and Indonesia. There for, Philippines also has enough data but raw data show the swing-type behavior so the third-order polynomial equation cannot fit well, while the other country have not enough data to conform our condition, 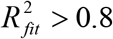. We find that Singapore having 2 values of as 1.54 (16 Feb to 37 March) and 6.22 (31 March-4 April). The higher are Indonesia (7.97), Singapore (6.22), Malaysia (3.86), Thailand (2.48), respectively. The higher *R*_0_ are corresponded to the higher total case as the total cases are found; Singapore (6588 cases), Indonesia (6575 cases), Malaysia (5384 cases), Thailand (2765 cases) (Worldmeter on 19 April 2020.). The more incubation period, the more value of are found.

After taking the regression method to raw data, we find that Malaysia, Philippines, Thailand, Brunei Darussalam, Viet Nam and Cambodia show the peak of the spreading. We take the extrapolate on equations to reach the end of time so “Bell-shape” occurred. To compare with the equal peak, we normalized the results with the maximum of each country and shown in Figure 1.

**Figure 1.**
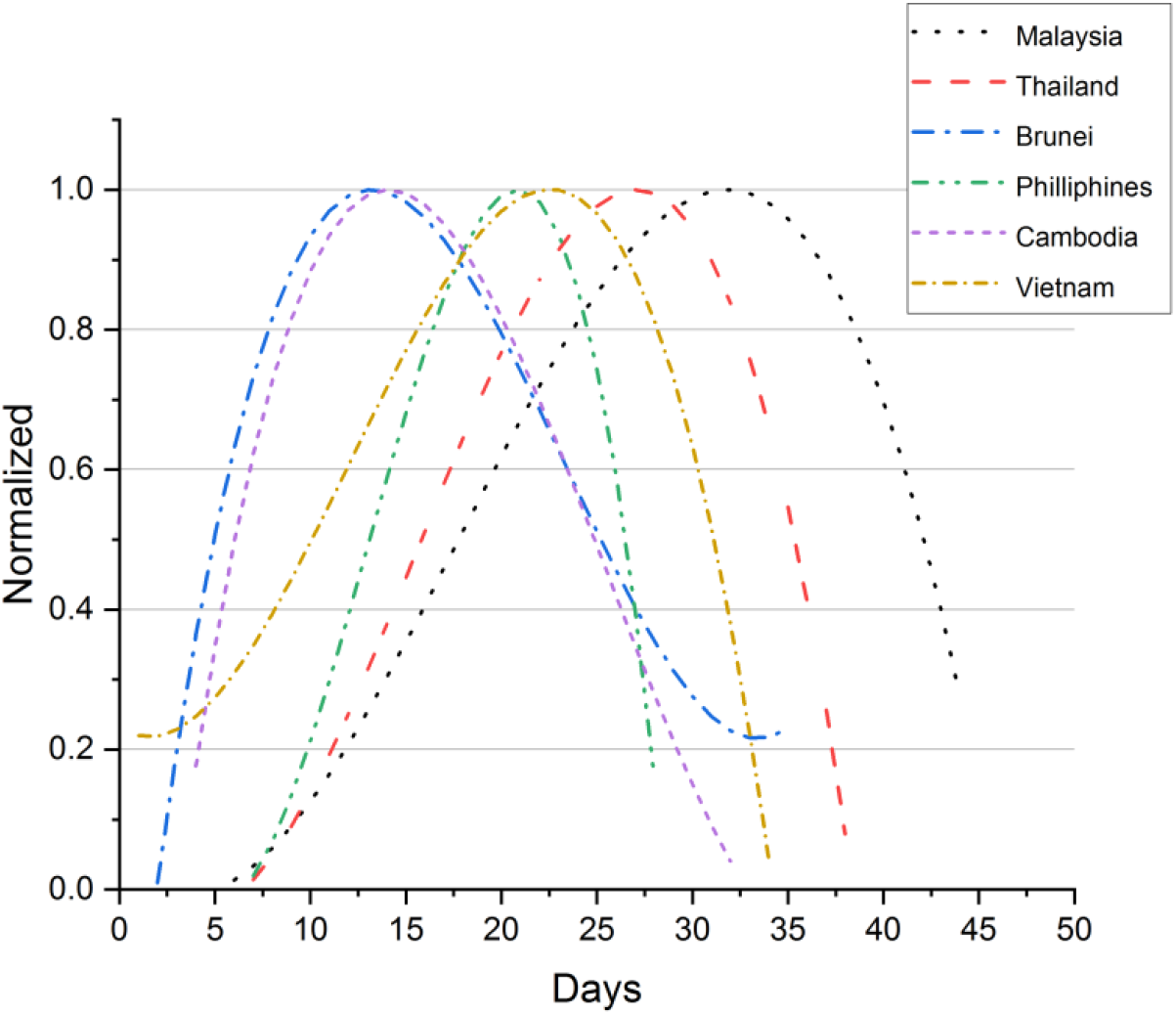
The normalized peak of spreading curve of an Asean.

Most of these countries found first-case on January; Thailand, Singapore, Viet Nam, Malaysia, Cambodia, Philippines and the other found on March; Indonesia, Brunei Darussalam, Lao PDR, Myanmar [Wikipedia]. It takes a time to be outbreak to global pandemic depending on condition of each country. In Figure. 1, the maximum of the spreading peak is found in chronological order as Brunei Darussalam, Cambodia, Philippines, Viet Nam, Thailand, and Malaysia, respectively. In this case Thailand and Malaysia have, 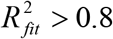, so the equation are well enough for consideration. The graph of Thailand has more slope than the graph of Malaysia and the total case of Malaysia (5384 cases) are higher than Thailand (2765 cases) (Worldometer on 19 April 2020.) [6]. This result is agree with more R_0_ of Malaysia than R_0_ of Thailand as Table 1.

The raw data of Indonesia and Singapore have shown that these countries take more time than the other country to get a peak. So, their curves do not show the Bell-shape at the time of consideration. However, we can calculate the R_0_ of them that are 6.22 and 7.97 for Singapore and Indonesia, respectively. The high value of the R_0_ mean that they will take more time to get a peak and the infective population will be abundant if government do nothing.

In this situation, the external stimulus is need for stimulate the system to exhibit the new character such as the FitzHugh - Nagumo equations [12,13]. The Eq.(5) is similar to the FitzHugh - Nagumo equations then we add the external stimulus, *I* _*ext*_ into Eq.(5),

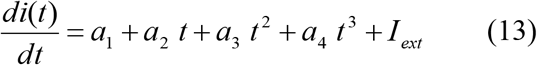

The external stimulus will reset the variable of system to become new value. If the external stimulus exceeds a certain threshold value, the system will generate a new behavior. In our model, we need the external stimulus to reduce the exponential increase to be the Bell-shape form. So, we set 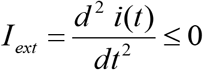, that is the condition for maximum value. The external stimulus is the policy of public-health organization of each country. The aim of public-health policy is to flatten the curve and decrease the maximum infected population of spreading over time. The basic protective measures against the Covid-19 such as wash your hands frequently and avoid touching eyes, nose and mouth (WHO). In Asean, aside from the basic suggestions from WHO, many policies are done for decrease the spreading of Covid-19. A common policy used are travel restriction, social distancing and work at home. The difference is the time of these policy beginning that depend on first case confirmed and the attention of government and people to solve this problem. The medical supply and living conditions are still the problem of this area [Wikipedia].

In this paper, we take the constant external stimulus as 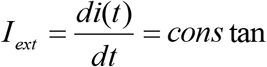. And this constant is equal to the latest value of daily infection per day.

From Figure 2 and Figure 3, Singapore need more the magnitude of external stimulus than Indonesia. However, the external stimulus for 14 days can stimulate to occur the peak of infective daily case of both country. So, the effective policy for decrease the spreading of Covid-19 should be at least 14 days. In Singapore the preventive measures were collectively called a ‘circuit breaker’. But there is still some unsuccessfulness of this policy. The small territory of this country makes some problems such as living conditions at foreign worker dormitories and social distancing on public transport is not long enough. In Indonesia, there are many policies were done by central government such as travel restrictions, making medical facility specially equipped to handle the coronavirus, social distancing, work from home and having stimulus policy on economic sector. [Wikipedia].

**Figure 2.**
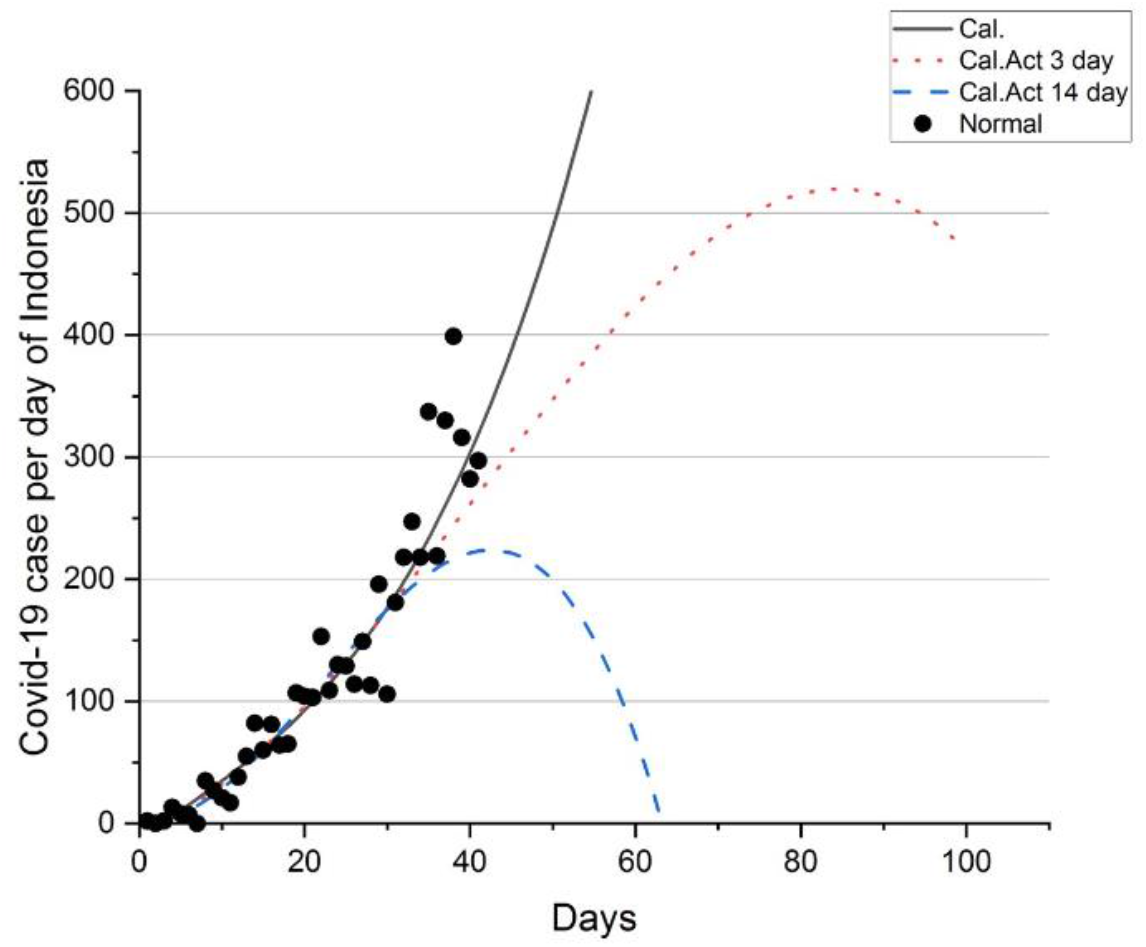
The dot “Normal” are raw data of daily case of Indonesia [6]. The “Cal” is from the regression. “Cal Act 3 day” is regression with the external stimulus for 3 days. “Cal Act 14 day” is regression with the external stimulus for 14 days.

**Figure 3.**
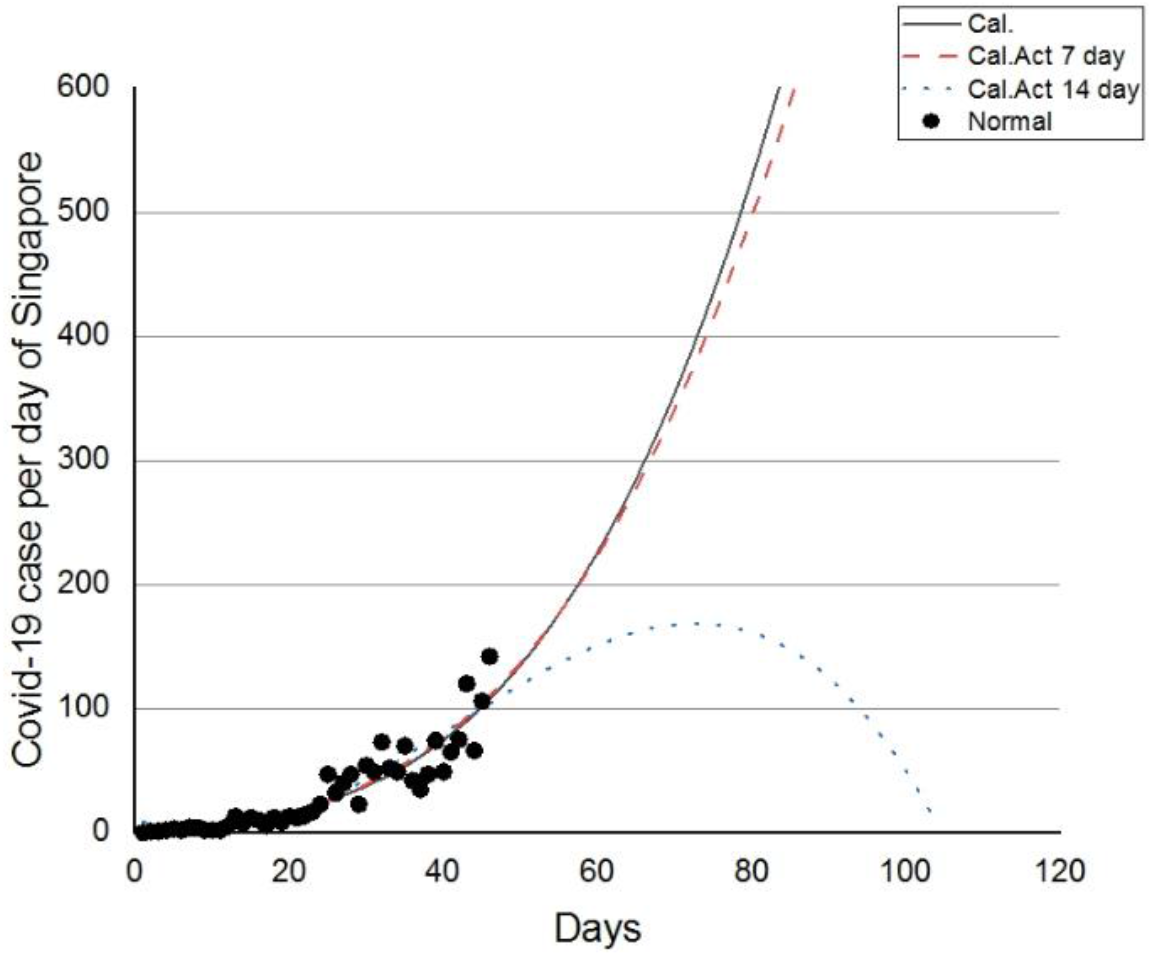
The dot “Normal” are raw data of daily case of Singapore [6]. The “Cal” is from the regression. “Cal Act 7 day” is regression with the external stimulus for 7 days. “Cal Act 14 day” is regression with the external stimulus for 14 days.

## 4. Conclusions

The Covid-19 raw data of infected rate of Asean are fitted with the regression method of third-order polynomial formula under the scope of SEIR model. We derived the reproductive index and set into 2 equations as 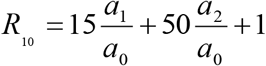 and 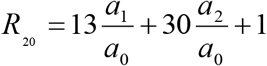 for the average incubation period as 5 days and 3 days with the average infection period as 10 day, respectively. The raw data show that there are only 4 country having enough data to fit well with third-order polynomial as Malaysia, Thailand, Singapore, and Indonesia. The higher *R*_0_ are Indonesia (7.97), Singapore (6.22), Malaysia (3.86), Thailand (2.48), respectively. And we also find that Singapore having 2 values of *R*_0_ as 1.54 (16 Feb to 37 March) and 6.22 (31 March-4 April). Our estimated R_0_ are in range that consistent with other preliminary estimates posted on public domains [14–17]. However, the peak of infected rate is not found for Singapore and Indonesia at the time of consideration. The model of external stimulus on raw data are added into polynomial equation of Singapore and Indonesia to control the Covid-19. We find that Singapore need more the magnitude of external stimulus than Indonesia. However, the external stimulus for 14 days can stimulate to occur the peak of infective daily case of both country.

## Data Availability

Worldometer from https://www.worldometers.info/coronavirus/

## Acknowledgments

We would like to thank to Srinakharinwirot University and Huachiew Chalermprakiet University for support at all the time of doing research.

